# Gut microbial metabolites lower 24-hour systolic blood pressure in untreated essential hypertensive patients

**DOI:** 10.1101/2022.06.20.22276673

**Authors:** Hamdi A. Jama, Dakota Rhys-Jones, Michael Nakai, Chu K Yao, Rachel E. Climie, Yusuke Sata, Dovile Anderson, Darren J. Creek, Geoffrey A. Head, David M. Kaye, Charles R. Mackay, Jane Muir, Francine Z. Marques

**Author notes:** **Corresponding author:** A/Prof Francine Marques, Hypertension Research Laboratory, School of Biological Sciences, Faculty of Science, Monash University, Melbourne, Australia.

## Abstract

**Background:** Fibres remain undigested until they reach the colon, where some are fermented by gut microbiota, producing metabolites called short-chain fatty acids (SCFAs). SCFAs lower blood pressure (BP) of experimental models, but their translational potential is unknown. We aimed to determine whether SCFAs lower 24-hour systolic BP (SBP) in untreated participants with essential hypertension.

**Methods:** We performed a phase II randomized placebo-controlled double-blind cross-over trial using SCFA-supplementation, delivered as acetylated and butyrylated high amylose maize starch (HAMSAB). Twenty treatment-naïve hypertensive participants were recruited from the community and randomised to 40g/day of HAMSAB or placebo. Participants completed each arm for three-weeks, with a three-week washout period between them. The primary endpoint was a 24-hour SBP decrease.

**Results:** Participants were on average 55.8±11.2-years old (mean±SD), had a body mass index (BMI) of 25.7±2.5km^2^/m, 30% were female, baseline 24-hour SBP 136±6mmHg. No adverse effects were reported. After the intervention, the placebo-subtracted reduction in 24-hour SBP was 6.1±9.9mmHg (*P*= 0.027). This was independent of age, sex, BMI and study arm. There was no statistical significance in the placebo arm. Day and night SBP were reduced by 6.5±12.3mmHg (*P*=0.01) and 5.7±9.8mmHg (*P*=0.02), respectively, and 24-h central SBP by 7.2±14.7 mmHg (*P*=0.005). HAMSAB increased levels of acetate and butyrate by 7.8-fold (*P*=0.016), shifted the microbial ecosystem, and expanded the prevalence of SCFA-producers.

**Conclusions:** We observed a clinically relevant reduction in 24-hour SBP in participants with essential hypertension treated with the gut microbial-derived metabolites acetate and butyrate. These metabolites may represent a novel option for lowering BP.

## Introduction

Uncontrolled high blood pressure (BP), also known as hypertension, is the primary cause of non-communicable diseases and global deaths.^1^ It is estimated that over 1 billion people worldwide have hypertension.^2^ The overwhelming majority of patients with hypertension remain with uncontrolled or suboptimal BP,^3^ which leads to cardiovascular disease (CVD).^1^ Western-style diets, which are low in fibre and high in saturated fats and sodium, raise the risk of developing hypertension and CVD.^4^ Conversely, those with a high intake of fruits and vegetables have lower BP.^5^ A recent systematic review and meta-analysis assessed the health impact of dietary fibre intake in 185 prospective studies and 58 clinical trials.^6^ Increased fibre intake, especially between 25 and 29 grams per day, led to a 15-30% reduction in all-cause and cardiovascular mortality and was associated with lower BP.^6^

Mounting evidence supports that gut dysbiosis is a common feature and contributor to experimental and clinical hypertension.^7,8^ Dietary fibre is defined as a carbohydrate that is not absorbed nor digested in the small intestine, thus reaching the large intestine undigested.^9^ Specific types of fibre are fermented by the gut microbiota, which produce metabolites called short-chain fatty acids (SCFAs), predominantly acetate, butyrate and propionate. These are energy sources for intestinal epithelial cells.^10^ Using experimental models, we discovered that the beneficial effects of dietary fibre to lowering BP and preventing CVD are due to SCFAs.^11^ These findings were subsequently validated in further experimental models,^12-14^ with acetate and butyrate having the most pronounced effects, lowering BP by 35% and 20%, respectively.^14^ However, there is no evidence that SCFAs may lower BP in hypertensive patients. A significant challenge in translating these findings has been the sustained delivery of SCFAs over a long period.^15^

We hypothesised that increased production of gut microbiota-derived metabolites acetate and butyrate reduce BP in untreated hypertensive patients. To overcome the challenge of delivering these SCFAs to the systemic circulation, we used high amylose maize starch (HAMS), a type of resistant starch II that can be acetylated (HAMSA) and butyrylated (HAMSB).^16^ Intestinal microbial fermentation of HAMSAB releases high levels of acetate and butyrate in the colon, which are subsequently absorbed and delivered to the systemic circulation.^17^ Thus, we aimed to conduct the first randomised clinical trial to determine if HAMSAB lowers BP in untreated hypertensive patients. We provide evidence that the SCFAs acetate and butyrate, delivered as HAMSAB, lowers systolic BP in humans and could be employed as a new BP-lowering strategy.

## Methods

### Study design

Human ethics approval was obtained from Monash University Human Research Ethics Committee (Study ID: 19203). The study was registered at the Australian New Zealand Clinical Trials Registry (ACTRN12619000916145) and followed the Declaration of Helsinki. All participants provided written informed consent before commencing the trial. A detailed protocol on the rationale and study design was previously published.^18^ Briefly, this study was a double-blind, randomised, placebo-controlled cross-over trial conducted in Melbourne, Australia.

### Participants

Recruitment started in July 2019 and finished in September 2021. Untreated hypertensive participants were recruited and screened for eligibility before being randomised. Inclusion criteria included being untreated for hypertension (as defined by the Australian National Heart Foundation guidelines, determined during the first study visit with 24-h systolic BP ≥130 mmHg and/or 24-h diastolic BP ≥80 mmHg), being of either sex (self-reported), 18-70 years of age, and having a body mass index (BMI) of 18.5–35 kg/m^2^. Exclusion criteria included any antihypertensive medication, an office BP measurement >165/100 mmHg, antibiotic treatment in the last three months, or probiotic intake over the previous six weeks. Those presenting with comorbidities (including type 1 or 2 diabetes or any gastrointestinal disease), pregnancy or specific dietary requirements (e.g. plant-based, gluten-free) or food intolerances were also excluded.

### Randomisation and masking

Study participants were randomised and stratified using REDCap software (Version 9.1.0 Tennessee) in a 1:1 ratio. This study was double-blinded; concealment of the allocated diet was labelling all food and material related to the trial as ‘Diet A’ and ‘Diet B’. The study coordinator was only unblinded after the trial was completed, when the analysis of the data was completed.

### Procedures

We worked with a research chef and dieticians to develop a suite of foods (described in ^18^) with similar appearance and taste that could be frozen and reheated and contained the placebo or HAMSAB. Following successful screening and randomisation, participants were assigned to Diet A or Diet B for three weeks. Diet A contained HAMSAB (obtained from Ingredion, USA) delivered as 40g/day divided into two portions of 20g in the morning meal and 20g in the evening meal, and Diet B contained 40g/day of placebo (corn starch or regular flour with no added resistant starches) delivered in a similar manner for three weeks. All other nutritional components (including energy, fat and protein) were the same. A complete breakdown of the nutritional content of the diets is given in Table S1 and S2. The three-week intervention was chosen based on a previously published paper.^14^ After a three-week washout period, participants were placed on the opposite arm for three weeks. All study food was stored in a –20°C freezer at Monash University. Participants were provided with a list of foods that were naturally high in resistant starches and SCFAs and were instructed to avoid them for the duration of the study. All participants were required to document dietary intake in a 3-day food diary immediately before commencement and at the end of each arm of the study for assessment of their habitual diet, and a daily food diary for the consumption of study meals for the duration of the study. Food diaries were then analysed for changes in dietary intake using the FoodWorks Professional software (Version 7.01, Xyris, Queensland) and resistant starch using an in-house database built into FoodWorks (J.M.). Adherence to the study diets were assessed as excellent using previously published criteria.^19^

### Blood pressure measurements

An average of three office BP measurements were taken by the trial coordinator using standard protocol under resting conditions (>5 min sitting with the researcher not in the room, seated, with back supported, and arms and legs uncrossed) using an automated digital BP monitor (Omron Healthcare, Japan, HEM-907). Participants were given a demonstration and detailed instruction following the Australian National Heart Foundation guidelines on performing at-home blood pressure monitoring using a calibrated and STRIDE approved BP monitor (Omron Healthcare, Japan, HEM-7121). Participants were instructed to take consecutive two measurements at the same time each day under rested conditions for the duration of the study. Participants were also fitted with an ambulatory blood pressure monitor (ABPM) (Mobil-O-Graph BP device, IEM Industrielle Entwicklung Medizintechnik) before and after the three-week dietary intervention, in a total of four visits. These devices measured BP every 15 minutes during the day and every 30 minutes at night. These measurements were used to confirm the office hypertensive diagnosis prior to enrolment in the trial. Participants were instructed to maintain a regular schedule and document any factors that may influence their BP readings (e.g. change in medication, stress or illness, bed time).

### Gastrointestinal transit and pH

Regional gastrointestinal transit times and pH were measured using the SmartPill™ Motility Testing System (Medtronic), as previously described.^20^ This was implemented later in the trial, thus performed only in a subset of participants (n=7 baseline, placebo and HAMSAB). After collecting blood samples, fasting participants were asked to eat a Smartbar and then swallow the SmartPill™ capsule (Medtronic), according to the manufacturer’s protocol. This allows real-time measurement, transmitted wirelessly to a wearable data receiver, of intraluminal pH through the whole gastrointestinal tract, as well as assessment of gastrointestinal transit. Participants fasted for 6 hours before resuming normal food and drink. Additionally, they had to always keep the receiver within 1.5 m of the body until the SmartPill capsule was passed. Data were then downloaded from the receiver on the SmartPill Motility Program and analysed for gastric emptying, small intestinal, colonic and whole gut transit time independently by two trained investigators (D.R.J., C.K.Y.). Any discrepancies were resolved via discussion and consensus between the two. As microbial fermentative activities differ across the different colonic regions, colonic pH profiles were further defined using median, minimum and maximum pH using previous protocol.^20^

### Short-chain fatty acids measurements

Fasting blood was collected in the morning, at least 12-hours after the last HAMSAB or placebo meal. Plasma SCFAS were quantified using mass spectrometry, as previously described.^21^ Briefly, 20µl of plasma was analysed in duplicates in a Q-Exactive Orbitrap mass spectrometer (Thermo Fisher Scientific) in conjunction with a Dionex Ultimate® 3000 RS high-performance liquid chromatography (HPLC) system (Thermo Fisher Scientific). We accepted a coefficient of variability <15%. Standard curves were constructed using the area ratio of the target analyte, and the internal standard in the range of each analyte was used. The levels of butyrate in some samples were below detection level of the standard curve; in this case they were considered half of the lowest measurable value, equivalent to 0.05 ng/ml.

### Faecal DNA extraction and 16S sequencing

Our study followed guidelines for faecal microbiota studies in hypertension,^22^ and the Strengthening The Organization and Reporting of Microbiome Studies (STORMS) reporting (Table S4).^23^ Stool samples were collected in tubes containing DNA/RNA Shield (Zymo Research) for microbial DNA extraction. Tubes were brought to the clinic immediately after a sample was produced, or were stored at –20°C for less than 24-hours and then brought to the clinic, where they were stored at –80°C until further processing. DNA was extracted using the DNeasy PowerSoil DNA isolation kit (Qiagen). The V4-V5 region of the bacterial 16S rRNA was amplified by PCR using the 515F and 926R primers (Bioneer) and was sequenced in an Illumina MiSeq sequencer (300 bp paired-end reads), as we recently described.^21^

### Bioinformatic analyses of the faecal microbiome

Sequence reads from samples were first analysed using the QIIME2 framework,^24^ as we previously described.^21^ Identification of changes in α-diversity (using indices Chao1, Shannon’s and Observed), β-diversity (Bray-Curtis index) and differentially abundant taxa analyses were performed on MicrobiomeAnalyst.^25^ We used edgeR to identify differential abundant taxa, and this was adjusted for multiple comparisons using a false discovery rate (FDR), where q<0.05 was considered significant.

### Inflammatory markers

High sensitivity Proquantum immunoassays for cytokines IL-10 (cat#A35590), IL1β (cat#A35574), IL17A (cat#A35611), IL6 (cat#A355735) were performed according to the manufacturer’s protocol in a QuantStudio 7 qPCR Instrument (Thermo Fisher Scientific) in duplicates from placebo and HAMSAB timepoints. The quantification was calculated based on the standard curve.

### Adverse effects

Adverse effects were closely monitored during the trial and discussed at each visit. Diets were well-tolerated, and no adverse effects were reported. We also used a 100mm Visual Analogue Scale to quantify gastrointestinal symptoms as previously utilised,^19^ where >30 mm out of a scale of 100 mm is considered a clinically significant increase in symptoms.

### Outcomes

The primary outcome of this study was a decrease in 24-hour systolic BP measured using ABPM measurements. Secondary outcomes included changes in plasma SCFA levels, circulating markers of inflammation and faecal microbial composition. We have further expanded changes to BP to analyse day, night and central systolic BP and home systolic BP.

### Sample size

In this proof-of-principle study, we estimated we would require 26 subjects to achieve 80% power with α=0.05 (calculated effect size 0.8) to determine a 7 mm Hg difference in 24-hour systolic BP after intake of modified HAMSAB for 3-weeks. We initially aimed to recruit 33 participants per group to allow a 20% dropout rate. Due to the SARS-COV-2 pandemic, recruitment was challenging and took longer than expected. As a result, the study stopped at the end of the allocated budget and staffing time. Thus, we stopped the trial at 21 participants recruited, with one dropout who started the trial on the Diet B (placebo) arm and BP reached >165/100 mmHg, requiring medication (Figure 1). Therefore, although we used a 1:1 randomisation, more participants were first allocated to Diet A than Diet B, and the calculated sample size was not reached.

**Figure 1.**
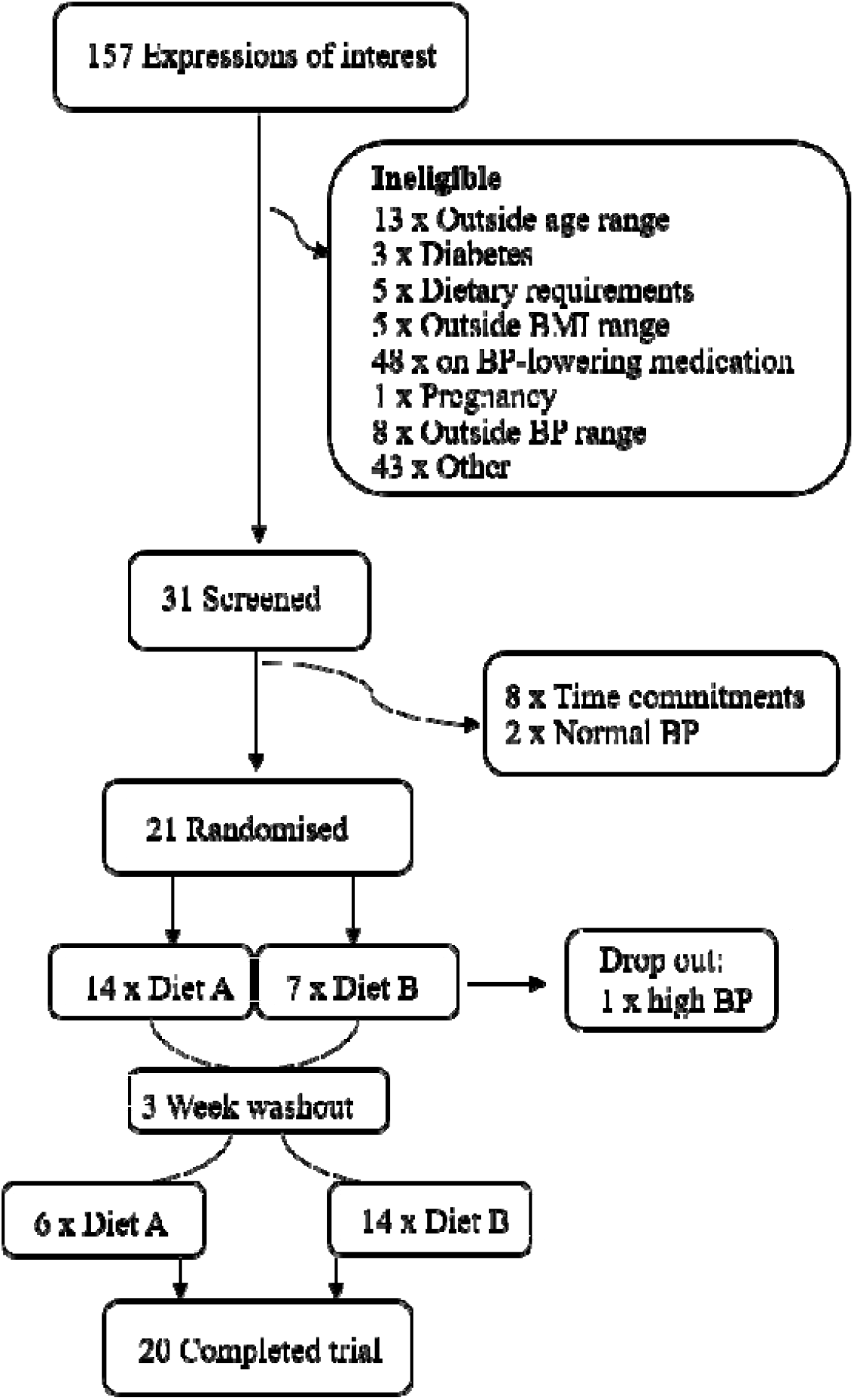
Recruitment summary. This study attracted 143 expressions of interest. 31 individuals met the inclusion criteria and were screened and of these 20 successfully completed the trial.

### Missing data

One participant randomised into Diet A first had to take antibiotics between their third and study final visit while they were on the Diet B (placebo). Thus, their last data point was imputed using the multiple imputation function in SPSS. We did not impute their data for the SmartPill data due to the small subsample size studied (n=7).

### Statistical analyses

Clinical data were analysed using SPSS software (version 25), while SCFAs, gastrointestinal pH and cytokine data were analysed in GraphPad Prism (version 6). Clinical data were checked for normality and then used paired two-tailed t-tests between baselines (Tables S4) and between each baseline and its intervention (placebo or HAMSAB). Sensitivity analyses adjusted by confounding variables were performed for 24-h systolic and diastolic BP, which were further analysed using a general linear mix (GLM) model with repeated measures between and within participants, adjusted for sex, BMI, age and study arm. These are shown as placebo-subtracted or baseline subtracted mean differences. All other data were analysed using GraphPad Prism (version 6) built-in paired two-tailed t-tests (with a confidence interval of the mean of differences shown as estimation plots) or one-way ANOVA with Benjamini and Hochberg’s false discovery rate adjustment for multiple comparisons (SmartPill data). All data are presented as mean±SEM or mean±SD, as detailed in figures and tables. *P*-value <0.05 was considered significant.

## Results

### Participant characteristics

Between 9 July 2019 and 28 August 2021, we received 157 expressions of interest and screened 31 participants. Of these, 21 were randomised to either the SCFAS-enriched (HAMSAB) or placebo (Figure 1). One participant dropped out of the study due to BP >165/100 mmHg before completion, having to receive medication. Therefore, 20 participants completed the trial. Baseline characteristics of the participants, including age, BMI, biochemical profile and BP, are shown in Table 1. The baseline mean±SD 24-hour systolic BP was 136±6mmHg and diastolic BP 87±7mmHg. Participants randomised to either the placebo or HAMSAB first had no significant differences in age, sex, BMI, office, and 24-h BP and biochemical profiles. However, participants randomised into the placebo arm had higher baseline non-HDL (*P*=0.02) and LDL cholesterol (*P*=0.04), and triglycerides (*P*=0.03) (Table S5). This resulted in an observed increase in LDL and non-HDL cholesterol and triglycerides with HAMSAB compared to its baseline (Table 2), as there was no difference between placebo and HAMSAB arms. Compared to baseline measurements, biochemical parameters remained unchanged in the placebo arm of the study, while there was a reduction in bilirubin in the HAMSAB arm (Table 2). All other parameters tested remained unchanged, including BMI (Table 2).

**Table 1.**
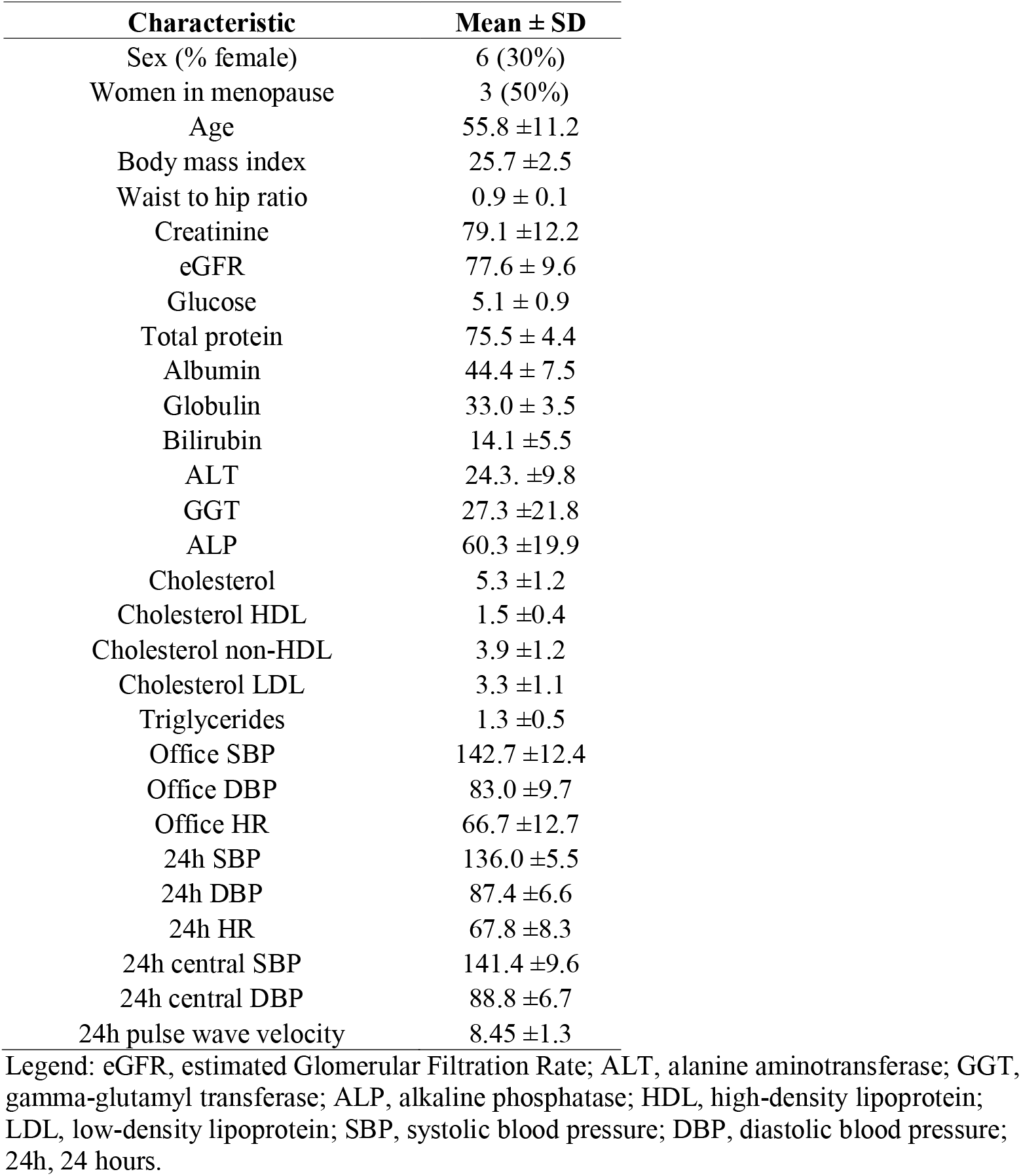
Baseline characteristics of the cohort.

| Characteristic | Mean ± SD |
| --- | --- |
| Sex (% female) | 6 (30%) |
| Women in menopause | 3 (50%) |
| Age | 55.8 ± 11.2 |
| Body mass index | 25.7 ± 2.5 |
| Waist to hip ratio | 0.9 ± 0.1 |
| Creatinine | 79.1 ± 12.2 |
| eGFR | 77.6 ± 9.6 |
| Glucose | 5.1 ± 0.9 |
| Total protein | 75.5 ± 4.4 |
| Albumin | 44.4 ± 7.5 |
| Globulin | 33.0 ± 3.5 |
| Bilirubin | 14.1 ± 5.5 |
| ALT | 24.3 ± 9.8 |
| GGT | 27.3 ± 21.8 |
| ALP | 60.3 ± 19.9 |
| Cholesterol | 5.3 ± 1.2 |
| Cholesterol HDL | 1.5 ± 0.4 |
| Cholesterol non-HDL | 3.9 ± 1.2 |
| Cholesterol LDL | 3.3 ± 1.1 |
| Triglycerides | 1.3 ± 0.5 |
| Office SBP | 142.7 ± 12.4 |
| Office DBP | 83.0 ± 9.7 |
| Office HR | 66.7 ± 12.7 |
| 24h SBP | 136.0 ± 5.5 |
| 24h DBP | 87.4 ± 6.6 |
| 24h HR | 67.8 ± 8.3 |
| 24h central SBP | 141.4 ± 9.6 |
| 24h central DBP | 88.8 ± 6.7 |
| 24h pulse wave velocity | 8.45 ± 1.3 |

Legend: eGFR, estimated Glomerular Filtration Rate; ALT, alanine aminotransferase; GGT, gamma-glutamyl transferase; ALP, alkaline phosphatase; HDL, high-density lipoprotein; LDL, low-density lipoprotein; SBP, systolic blood pressure; DBP, diastolic blood pressure; 24h, 24 hours.

**Table 2.**
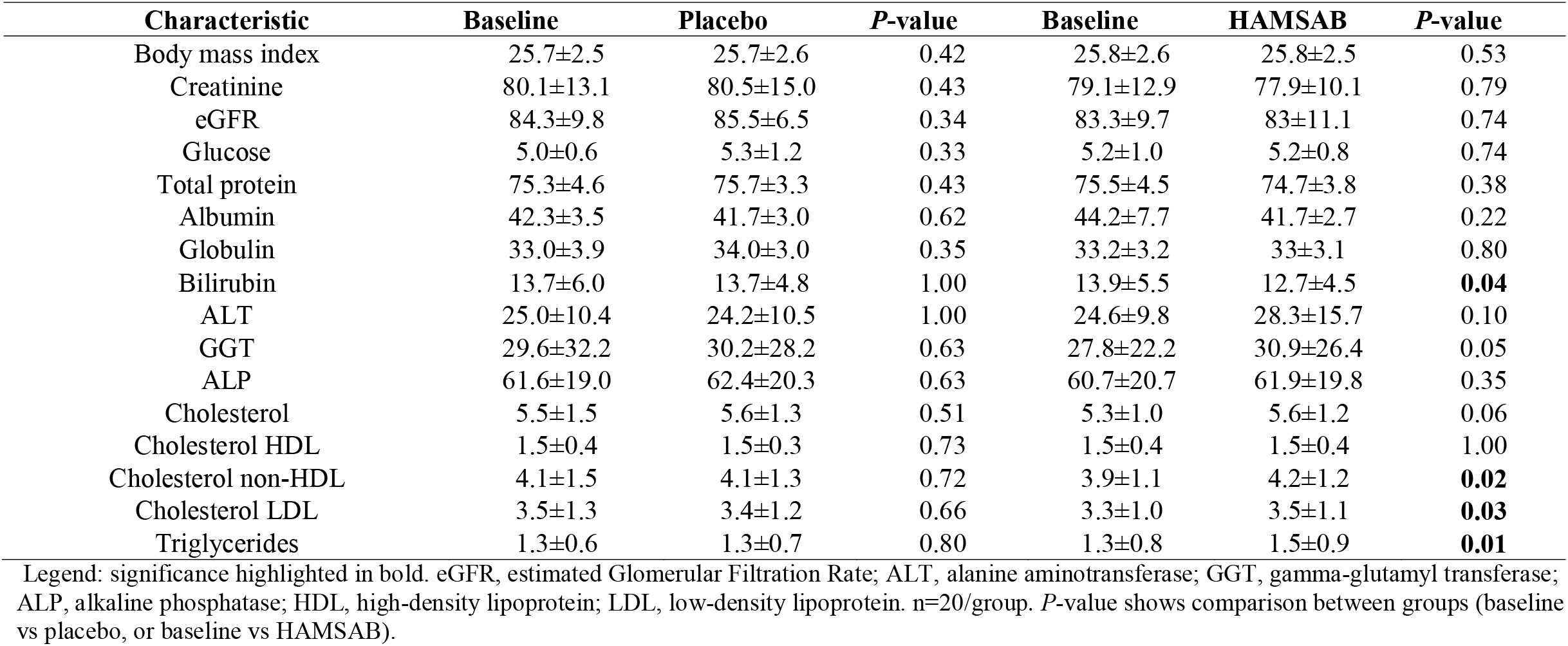
Comparison of changes in biochemical parameters from baseline in the placebo and HAMSAB diet arm.

### Dietary adherence and gastrointestinal tolerance

HAMSAB and placebo diets were matched in macronutrient content, including protein, fat, and carbohydrates (Table S1). The only difference was in overall dietary fibre (placebo 4.6g vs HAMSAB 19.7g) and resistant starch content (placebo 0.4g vs HAMSAB 15.8g). The HAMSAB and placebo diets were well tolerated, with participants consuming 93% of the meals provided. Analysis of food diaries showed that there was no difference in nutritional intake specifically protein, total fat, sodium and potassium between visits (Table S2). The only difference observed was an increase in fibre (+40%, *P*=0.0003) and resistant starch (566%, P<0.0001) between baseline and HAMSAB (Table S2), as expected. Participants in the HAMSAB arm scored higher for abdominal pain (*P*=0.024), while in the placebo arm they scored lower symptoms of bloating (*P*=0.022) in the VAS scale (Table S3).

### Blood pressure

We observed no differences in systolic BP in the placebo arm (Table 3 and S5). We observed a significant decrease in 24-hour systolic BP (baseline-to-HAMSAB: *P*=0.03, Table 3), with a placebo-subtracted mean difference of –6.1 mmHg (95% CI: –1.4-10.7mmHg *P*<0.0001, Figure 2a-b). We validated these using a GLM model, adjusted for sex, BMI, age and study arm, with baseline-to-HAMSAB having a decrease of 4.1 mmHg (*P*=0.027). In comparison, baseline-to-placebo had no difference (+2 mmHg, *P*=0.176). We also observed a significant decrease in both day and night systolic BP (respective placebo-subtracted mean difference: – 6.4[95% CI: –0.7-12.2mmHg], *P*=0.01; –6.32 [–1.1-10.3mmHg], *P*=0.02; Table S6), and central 24-h systolic BP (placebo-subtracted mean difference: –7.3 mmHg[–1.6-16.2mmHg], *P*=0.005, Table 3). We then examined changes to 24-h diastolic BP and observed no statistically significant change with HAMSAB (baseline-to-HAMSAB difference: *P*=0.23, Table 3 and Table S6), with a placebo-subtracted mean difference of –1.7 mmHg (*P*=0.179, Figures 2c-d). In the placebo group, there was an increase in 24-h diastolic BP (+3.84 mmHg, *P*<0.0001, Table 3). Interestingly, we did not observe a change in stroke volume, cardiac output or heart rate; however, HAMSAB borderline significantly reduced total vascular resistance (*P*=0.049, Table 3).

**Table 3.**
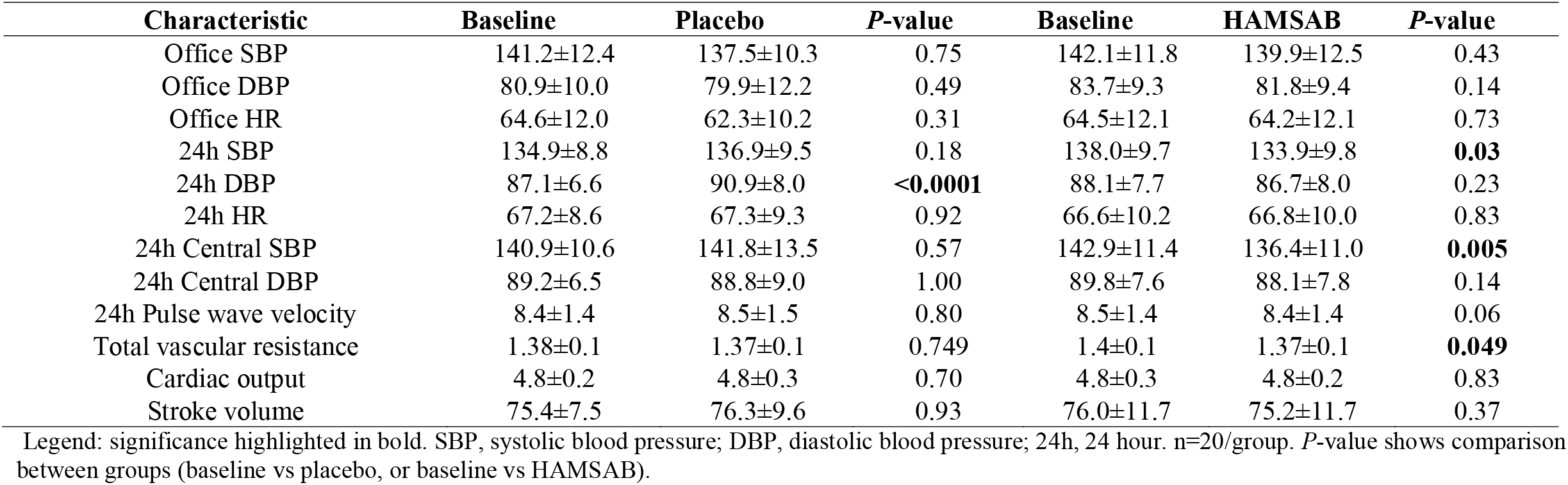
Comparison of changes in BP parameters from baseline in the placebo and HAMSAB diet arm.

| Characteristic | Baseline | Placebo | *P*-value | Baseline | HAMSAB | *P*-value |
| --- | --- | --- | --- | --- | --- | --- |
| Office SBP | 141.2±12.4 | 137.5±10.3 | 0.75 | 142.1±11.8 | 139.9±12.5 | 0.43 |
| Office DBP | 80.9±10.0 | 79.9±12.2 | 0.49 | 83.7±9.3 | 81.8±9.4 | 0.14 |
| Office HR | 64.6±12.0 | 62.3±10.2 | 0.31 | 64.5±12.1 | 64.2±12.1 | 0.73 |
| 24h SBP | 134.9±8.8 | 136.9±9.5 | 0.18 | 138.0±9.7 | 133.9±9.8 | **0.03** |
| 24h DBP | 87.1±6.6 | 90.9±8.0 | **<0.0001** | 88.1±7.7 | 86.7±8.0 | 0.23 |
| 24h HR | 67.2±8.6 | 67.3±9.3 | 0.92 | 66.6±10.2 | 66.8±10.0 | 0.83 |
| 24h Central SBP | 140.9±10.6 | 141.8±13.5 | 0.57 | 142.9±11.4 | 136.4±11.0 | **0.005** |
| 24h Central DBP | 89.2±6.5 | 88.8±9.0 | 1.00 | 89.8±7.6 | 88.1±7.8 | 0.14 |
| 24h Pulse wave velocity | 8.4±1.4 | 8.5±1.5 | 0.80 | 8.5±1.4 | 8.4±1.4 | 0.06 |
| Total vascular resistance | 1.38±0.1 | 1.37±0.1 | 0.749 | 1.4±0.1 | 1.37±0.1 | **0.049** |
| Cardiac output | 4.8±0.2 | 4.8±0.3 | 0.70 | 4.8±0.3 | 4.8±0.2 | 0.83 |
| Stroke volume | 75.4±7.5 | 76.3±9.6 | 0.93 | 76.0±11.7 | 75.2±11.7 | 0.37 |

Legend: significance highlighted in bold. SBP, systolic blood pressure; DBP, diastolic blood pressure; 24h, 24 hour. n=20/group. *P*-value shows comparison between groups (baseline vs placebo, or baseline vs HAMSAB).

**Figure 2:**
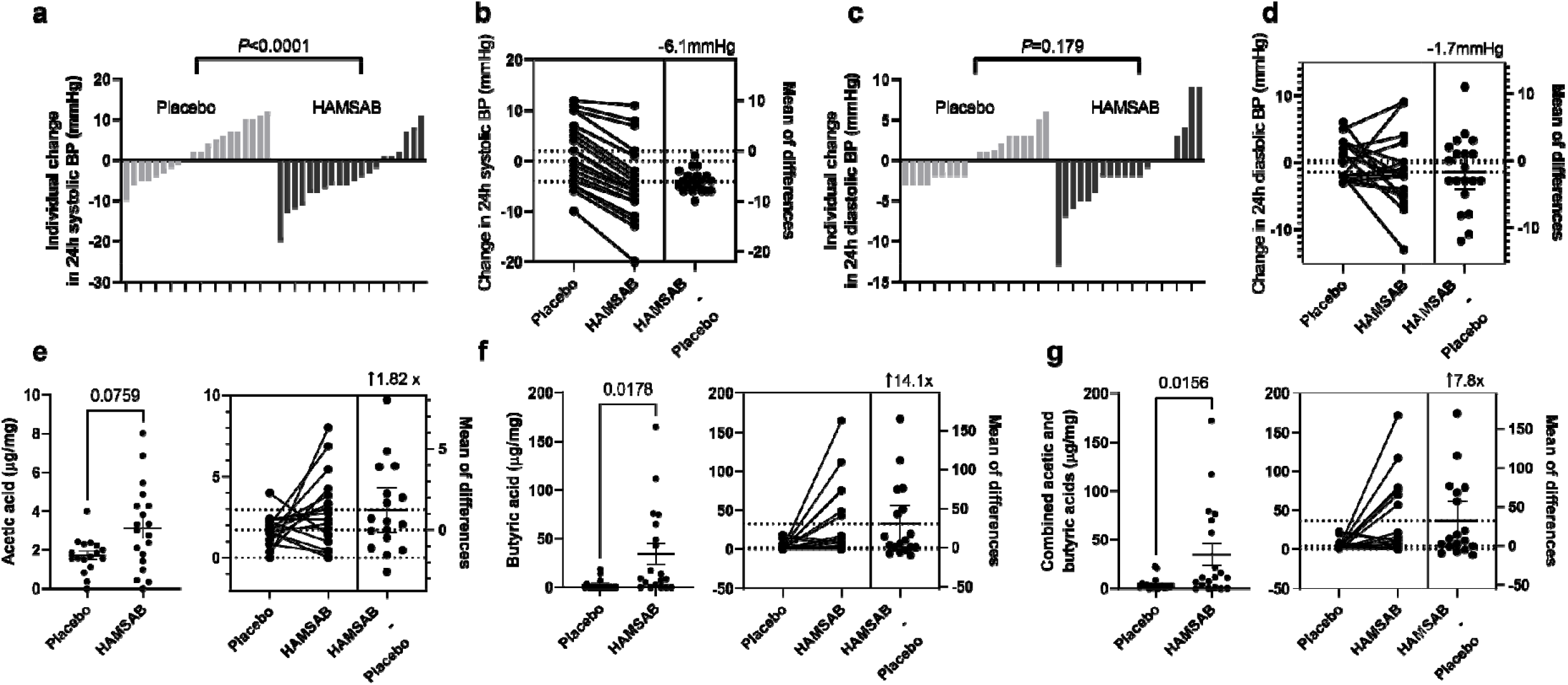
HAMSAB diet reduces in 24-h ambulatory systolic blood pressure (BP) and increases plasma short-chain fatty acids. a) Individual changes in 24-h systolic BP and b) mean change in 24-h systolic BP between placebo and HAMSAB and placebo subtracted mean difference. c) Individual change in 24-h diastolic BP and d) mean change in 24-h diastolic BP between placebo and HAMSAB and placebo subtracted mean difference. Plasma concentrations of e) acetic acid, f) butyric acid, g) combined total acetic acid and butyric acid. n=20/treatment group. Error bars represent ±SEM.

We did not observe a change in office systolic or diastolic BP (Table 2). However, similarly to ABPM, home systolic BP decreased over the 21-day intervention with the HAMSAB diet, particularly around day 9 and day 14 after the intervention started (Figure S1a). Participants on the HAMSAB diet achieved a –4.6-mmHg reduction in systolic BP when we compared the first three days of the intervention to the last three days (*P*=0.117, Figure S1b). Moreover, when we compared the first three days to day 14, systolic BP dropped by –4.8 mmHg (*P*=0.029, Figure S1c). Conversely, the participants on the placebo had no difference in systolic BP through the intervention (Figure S1d-e). Overall, the placebo-subtracted mean difference in home systolic BP was –1.48 mmHg (*P*=0.001, Figure S1g). Participants on the HAMSAB diet also had a modest non-significant drop in diastolic BP (Figure S1h-i) with a –2.4 mmHg decrease at day 14 (*P*=0.055) and no change in home diastolic BP in the placebo arm (Figure S1k-m). The placebo-subtracted mean difference in home diastolic BP was –0.79 mmHg (*P*=0.0295, Figure S1n). Together, this data supports that HAMSAB successfully reduced 24-h and home systolic BP. A larger number of participants may be needed to achieve statistical significance in diastolic BP changes.

### Plasma short-chain fatty acids

Compared to the placebo intervention, HAMSAB increased circulating acetate by 1.8-fold, but it did not reach statistical significance (*P*=0.076, Figure 2e). This may be explained by the bloods being collected more than 12-hours after the last HAMSAB meal. HAMSAB significantly increased the levels of butyrate 14-fold (*P*=0.0178, Figure 2f), as well as the combined plasma levels of acetate and butyrate by 7.8-fold compared to the placebo (*P*=0.0156, Figure 2g). These data confirm that HAMSAB delivers systemically high levels of SCFAs.

### Gastrointestinal pH and transit

We characterised the whole gastrointestinal transit and pH using real-time tracking in a subset of the cohort. We found no significant differences in regional transit times when comparing baseline, HAMSAB and placebo diet (Figure S2a-d). The colon is susceptible to changes in pH due to microbial communities releasing acidic metabolic by-products such as SCFAS, thus, providing an *in vivo* quantification of SCFAs. Given that HAMSAB delivers high levels of SCFAS, we wanted to determine if this would be reflected in colonic pH. We observed no difference in colonic minimum and median pH (Figure S2e-f). However, there was a significant decrease in maximum colonic pH on the HAMSAB diet compared to baseline (Figure S2i), with a significant reduction in the baseline-subtracted mean difference in pH in the HAMSAB arm (–0.45, *P*=0.033, Figure S2h).

### Faecal microbiome

We first compared the baseline faecal microbiome before starting the trial and after the three-week washout period. We observed no significant difference in microbial α-(Chao1, *P=*0.594, observed species, *P=*0.594, Shannon, *P*=0.870) and β-diversity (Bray-Curtis index, *P=*0.952) (Figure 3a-b). These data suggest the restoration of baseline microbiota was achieved during the washout period. Next, we compared the faecal microbiota composition of the HAMSAB diet and placebo. HAMSAB supplementation significantly decreased species richness (Chao1 *P*=0.009, observed species *P*=0.009), but not evenness (Shannon, *P*=0.117) (Figure 3c). Moreover, three weeks on the HAMSAB diet was sufficient to shift the β-diversity (Bray-Curtis index, *P=*0.037; Figure 3d). HAMSAB significantly increased the relative abundance of 6 taxonomic features (Table S7), including 2 SCFAS-producers, *Parabacteroides distasonis* (log2FC=2.8, FDR q-value=0.003) and *Ruminococcus gauvreauii* (log2FC=3.5, FDR q-value=0.0029) compared to placebo (Figure 3e-f). These findings support that HAMSAB treatment fosters the expansion of microbes that produce SCFAS in conjunction with releasing high levels of the pre-conjugated acetate and butyrate.

**Figure 3:**
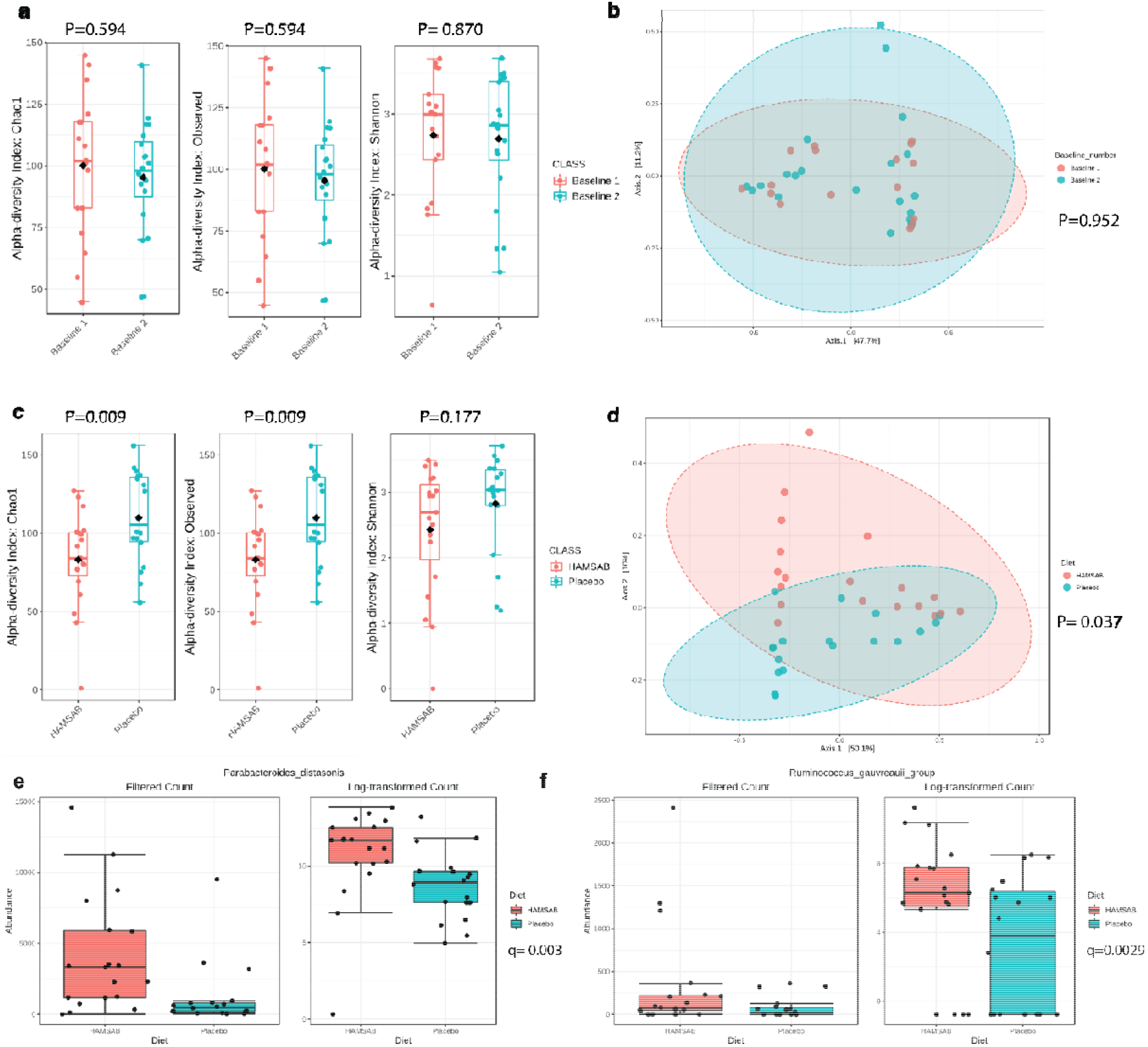
HAMSAB promotes the expansion of SCFA-producing commensal bacteria. 16S rRNA was sequences were obtained from faecal samples and analysed using QIIME2. a) Chao1, observed and Shannon’s alpha diversity indices analysed using Microbiome analyst software comparing baseline 1 and baseline 2. b) Principal coordinate analysis plot showing weighted Bray-Curtis index of at baseline of participants randomised to HAMSAB or placebo diet. c) Chao1, observed and Shannon’s alpha diversity indices comparing HAMSAB and placebo diet d) Principal coordinate analysis plot showing beta diversity measured by Bray-Curtis index of baselines of participants after HAMSAB or placebo diet. Filtered count and log-transformed count showing relative abundances of e) *Parabacteroides distasonis* and f) R*uminococcus gauvreauii*. n=19-20/treatment group.

### Plasma cytokines

SCFAs are thought to have anti-inflammatory properties,^10^ while hypertension is associated with increased pro-inflammatory markers, including IL17A, IL1b and IL6, and reduced anti-inflammatory marker IL10.^26^ Thus, we used a highly sensitive immunoassay to quantify the levels of plasma IL17A, IL1b, IL10 and IL6 between the HAMSAB and placebo treatment groups. We found no statistically significant difference in these cytokines (IL10: *P=*0.47; IL6: *P=*0.46, Il1β: *P=*0.40, IL17A: *P=*0.38, Figure S3), suggesting that three-weeks on the HAMSAB intervention did not impact plasma cytokine levels.

## Discussion

Adequate BP control remains poorly achieved globally, contributing to the rise in cardiovascular death.^2^ In recent years, SCFAs have emerged as potential therapeutic agents to treat hypertension in experimental studies.^11,14^ We investigated if a three-week intervention with HAMSAB, a specialised high-fibre supplement delivering high quantities of the SCFAs acetate and butyrate, can lower BP. To our knowledge, this is the first clinical evidence that delivering SCFAs lowers home, 24-h and central systolic BP in essential hypertensive patients, independently of sex, age, and BMI. HAMSAB diet also modulated the faecal microbiome, increasing SCFA-producers and raising SCFAs levels in the systemic circulation. As a result, this proof-of-concept study demonstrates the feasibility of employing supplementation to deliver high doses of microbial-derived metabolites to lower BP in essential hypertension. The reduction in systolic BP observed was equivalent to that achieved with conventional antihypertensive mono-treatment,^27^ and is calculated to reduce coronary death by 9% and stroke death by 14%.^28^

SCFAs are metabolites produced by commensal gut microbiota.^10^ These metabolites can be absorbed into the bloodstream and affect the host’s physiology.^10^ Current evidence suggests that the SCFAs orchestrate several complex signalling pathways that alter systemic transcriptional regulation^11^ and activate SCFA-receptors, such as GPR43.^14^ In experimental models, these culminate in reduced inflammation, lower BP, and prevention of hypertensive-induced end-organ damage.^11,14^ Our findings add to this growing data by showing that HAMSAB increases levels of acetate and butyrate in the plasma and that they indeed have BP-lowering effects in humans. This effect was larger than that reported for dietary fibre interventions alone.^6^ This finding is important because hypertension is associated with decreased circulating SCFAs^29^ and reduced SCFA-sensing receptor *GPR43*.^30^ While we did not measure SCFAS levels in the faeces, our data from *in vivo* real-time monitoring using the SmartPill supports that HAMSAB reduced the maximum pH in the colon. This is reflective of the distal colon, and suggests an increase in fermentation of dietary fibre in this area where carbohydrate fermentation rates are usually low due to depletion of fibrous substrates.

Increasing evidence supports the role of the gut microbiome in BP regulation.^7^ Dysbiosis, or functional and compositional abnormalities in the gut microbiota, have been described in clinical and experimental hypertension.^7,11,14,29,30^ Evidence supports the hypertensive gut microbiota has a depletion of SCFAs-producing bacteria.^29,30^ Subsequently, this depletion may impair SCFA-dependent pathways necessary for adequate BP control.

Intervention with HAMSAB supported the expansion of SCFA-producing commensal microbes *Parabacteroides distasonis* and *Ruminococcus gauvreauii* and supported the restoration of local production of SCFAs by these microbes. Belonging to the Lachnospiraceae family, decreased abundance of *Ruminococcus gauvreauii* is associated with lower levels of SCFAs and poorer cardiovascular outcomes.^31,32^ *Ruminococcus* sp. has a lower prevalence in untreated hypertensive^30^ and heart failure patients,^33^ and is negatively associated with 24-h systolic BP.^30^ Similarly, *Parabacteroides distasonis* is commensal bacteria that metabolises carbohydrates in the colon and mediates local inflammation.^34^ *Parabacteroides* genus is considered a member of the 20 core microbes in the human gut.^35^ *Parabacteroides distasonis* ferments dietary oligosaccharides to produce acetate and propionate.^17^ Our findings suggest that HAMSAB modulated systolic BP by increasing SCFA production and prevalence of SCFA-producers such as *Ruminococcus* sp and *Parabacteroides* sp. Further studies will need to determine if these taxa are required for the systolic BP-lowering effect we observed.

Contrary to hypothesised, we did not observe a reduction in cytokines with HAMSAB intake. These findings are consistent with a 5-day high fibre intervention, which did not change the frequencies of T cells in healthy participants.^15^ However, although of borderline significance, HAMSAB reduced another well-known factor that regulates BP: total peripheral resistance. This is consistent with findings in experimental hypertension.^14^ Vascular tone is an essential contributor to total vascular resistance. Early studies in rat and human arteries showed acetate and butyrate have a non-specific vasorelaxation effect.^36,37^ The mechanism was independent of traditional BP-lowering effects including endothelial and sympathetic activation.^36,37^ Given the large quantities of acetate and butyrate delivered through HAMSAB, it is possible that these metabolites influence vascular tone, resulting in lower total peripheral resistance. Further studies are needed to determine the specific mechanisms involved.

The main limitations of our study are the small sample size and lack of long-term follow-up. To adequately conclude the benefits of supplementary delivery of SCFAS to control BP, larger and longer-term trials need to be conducted. While our study detected significant changes in systolic BP, a larger sample size could yield more conclusive results for other variables such as diastolic BP. However, recruiting untreated hypertensive patients during the pandemic was a significant barrier, as most appointments were being carried out via telehealth, where BP was not measured. Our trial also had strengths, including the cross-over design, the double-blinding (which is rare in dietary studies), the use of HAMSAB to deliver high levels of SCFAs, and the use of ABPM and home BP monitoring.

In conclusion, our study provides the first evidence that delivering high levels of the microbial-metabolites acetate and butyrate reduces systolic BP and total vascular resistance in essential hypertensive patients. Furthermore, HAMSAB enriched essential SCFAs-producing microbes, which compounds the overall levels of SCFAs. Consumption of SCFAs directly, via the use of HAMSAB or other strategies, may represent a novel therapeutic option for hypertensive patients.

## Data Availability

The microbiome data described in this article will be made available at the GenBank Nucleotide Database (under submission). The other data underlying this article will be shared upon reasonable request to the corresponding author.

## Contributors

FZM, DMK, CRM and JM conceived and designed the study. HAJ wrote the first draft of the report with input from FZM; both HAJ and FZM accessed and verified the data, and did the statistical analyses. DRJ (managed by FZM and JM) coordinated the trial, recruited participants, and collected samples and data. YS helped with recruitment. MN (microbiome), REC and GAH (blood pressure), DA and DJC (metabolites), HAJ (cytokines) contributed with methods. All authors had full access to all data in the study, revised the manuscript critically, approved the version to be published, and had final responsibility for the decision to submit for publication.

## Acknowledgements

We would like to acknowledge the Monash Proteomics and Metabolomics Facility for SCFAS analysis and the Monash Bioinformatics Platform for access to M3 servers. We also would like to acknowledge Professor Markus Schlaich and Jonathon Sesa-Ashton for their help with the recruitment and Trish Veitch for her help developing the recipes used in the trial.

