## Supplemental file for "Gut microbial metabolites lower 24-hour systolic blood pressure in untreated essential hypertensive patients"

**Running title:** Gut microbial metabolites lower human blood pressure

Hamdi A. Jama[1], Dakota Rhys-Jones[1,2], Michael Nakai[1], Chu K Yao[2], Rachel E. Climie[3], Yusuke Sata[4], Dovile Anderson[5], Darren J. Creek[5], Geoffrey A. Head[4], David M. Kaye[6,7,8], Charles R. Mackay[9], Jane Muir[2], Francine Z. Marques[1,6]*

[1]Hypertension Laboratory, School of Biological Sciences, Faculty of Sciences, Monash University, Clayton, Victoria, Australia; [2]Department of Gastroenterology, Central Clinical School, Monash University, Melbourne, Victoria, Australia; [3]Menzies Institute for Medical Research, University of Tasmania, Hobart, Tasmania, Australia; [4]Neuropharmacology Laboratory, Baker Heart and Diabetes Institute, Melbourne, Victoria, Australia; [5]Monash Institute of Pharmaceutical Sciences, Monash University, Melbourne, Australia; [6]Heart Failure Research Group, Baker Heart and Diabetes Institute, Melbourne, Australia; [7]Department of Cardiology, Alfred Hospital, Melbourne, Australia; [8]Central Clinical School, Faculty of Medicine Nursing and Health Sciences, Monash University, Melbourne, Australia; [9]Department of Microbiology, Biomedical Discovery Institute, Faculty of Medicine, Nursing and Health, Monash University, Clayton, Victoria, Australia.

**Table S1.** Summary of the composition of the placebo and HAMSAB interventions per food item provided to the participants. One food item was consumed in the morning and one in the evening.

| Nutrient | Placebo | HAMSAB |
|---|---|---|
| **Energy (kJ)** | 3042 | 2999 |
| **Fat (g)** | 17.2 | 18.2 |
| **Carbohydrate (g)** | 42.6 | 41.1 |
| **Protein (g)** | 17.3 | 17.5 |
| **Dietary fibre (g)** | 4.6 | 19.7 |
| **Resistant starch (g)** | 0.4 | 15.8 |
| **Potassium (mg)** | 431.7 | 444.3 |
| **Sodium (mg)** | 523.0 | 552.0 |

**Table S2.** Dietary intake of participants during the trial, measured by 3-day food diaries.

| **Nutrient** | **Baseline** | **Placebo** | ***P-value*** | **Baseline** | ***HAMSAB*** | ***P-value*** |
|---|---|---|---|---|---|---|
| **Energy (kJ)** | 8632 ±1663 | 9494 ±2389 | 0.52 | 8185 ±1700 | 9303 ±1633 | 0.25 |
| **Carbohydrates (g)** | 202 ±56 | 214±63 | 0.59 | 200 ±57 | 234±41 | 0.30 |
| **Protein (g)** | 93±23 | 94 ±22 | >0.9999 | 82±19 | 89 ±16 | 0.70 |
| **Total fat (g)** | 87 ±26 | 99 ±30 | 0.48 | 82 ±20 | 93 ±20 | 0.43 |
| **Saturated fat (g)** | 31 ±11 | 37±10 | 0.35 | 31 ±9 | 33 ±8 | 0.91 |
| **Dietary-fibre (g)** | 27 ±6 | 22 ±7 | 0.00 | 22 ±9 | 38 ±6 | **<0.0001** |
| **Resistant-starch (g)** | 3 ±1 | 3 ±1 | 0.99 | 3 ±1 | 17 ±2 | **<0.0001** |
| **Sodium (mg)** | 2445 ±1109 | 2666 ±678 | 0.84 | 2445 ±683 | 2507 ±499 | 0.73 |
| **Potassium (mg)** | 3647 ±708 | 3351 ±944 | 0.68 | 2995 ±780 | 3382 ±679 | 0.42 |

Legend: significance highlighted in bold. Data shown as mean ± SD. n=20/group.

**Table S3.** Strengthening The Organization and Reporting of Microbiome Studies (STORMS) reporting checklist.

| Number | Item | Yes/No/NA | Comments or location in manuscript |
|---|---|---|---|
| **Abstract** | | | |
| 1 | Structured or Unstructured Abstract | Yes | Page 2 |
| 1.1 | Study Design | Yes | Page 2 |
| 1.2 | Sequencing methods | Yes | Page 2 |
| 1.3 | Specimens | | |
| **Introduction** | | | |
| 2 | Background and Rationale | Yes | Page 4 |
| 2.1 | Hypotheses | Yes | Page 4 |
| **Methods** | | | |
| 3 | Study Design | Yes | Page 5 |
| 3.1 | Participants | Yes | Page 5 |
| 3.2 | Geographic location | Yes | Page 5 |
| 3.3 | Relevant Dates | Yes | Page 5 |
| 3.4 | Eligibility criteria | Yes | Page 5-6 |
| 3.5 | Antibiotics Usage | Yes | Page 6 |
| 3.6 | Analytic sample size | Yes | Page 10 |
| 3.7 | Longitudinal Studies | NA | Randomised clinical trial, samples were collected across 4 timepoints |
| 3.8 | Matching | NA | Crossover study |
| 3.9 | Ethics | Yes | Page 5 |
| 4 | Laboratory methods | Yes | Page 8 |
| 4.1 | Specimen collection | Yes | Page 8 |
| 4.2 | Shipping | Yes | Page 8 |
| 4.3 | Storage | Yes | Page 8 |
| 4.4 | DNA extraction | Yes | Page 8 |

| | | | |
|---|---|---|---|
| 4.5 | Human DNA sequence depletion or microbial DNA enrichment | NA | |
| 4.6 | Primer selection | Yes | Page 8 |
| 4.7 | Positive Controls | Yes | Page 9 |
| 4.8 | Negative Controls | Yes | Page 8 |
| 4.9 | Contaminant mitigation and identification | Yes | Page 8 |
| 4.1 | Replication | NA | |
| 4.11 | Sequencing strategy | Yes | Page 8 |
| 4.12 | Sequencing methods | Yes | Page 8 |
| 4.13 | Batch effects | No | Some samples were sequenced in a different batch, however PCoA analyses showed no difference between batches |
| 4.14 | Metatranscriptomics | NA | |
| 4.15 | Metaproteomics | NA | |
| 4.16 | Metabolomics | Yes | Page 8 |
| 5 | Data sources/ measurement | Yes | Page 8 |
| 6 | Research design for causal inference | Yes | Page 5 |
| 6.1 | Selection bias | NA | |
| 7 | Bioinformatic and Statistical Methods | Yes | Page 9 |
| 7.1 | Quality Control | Yes | Page |
| 7.2 | Sequence analysis | Yes | Page 9 |
| 7.3 | Statistical methods | Yes | Page 9 |
| 7.4 | Longitudinal analysis | NA | |
| 7.5 | Subgroup analysis | NA | |
| 7.6 | Missing data | NA | |
| 7.7 | Sensitivity analyses | NA | |
| 7.8 | Findings | Yes | Page |

| | | | |
|---|---|---|---|
| 7.9 | Software | Yes | Page 9 |
| 8 | Reproducible research | Yes | Page |
| 8.1 | Raw data access | NA | Data will be uploaded to NCBI Sequence Read Archive |
| 8.2 | Processed data access | No | We will provided access to all raw data. |
| 8.3 | Participant data access | No | Individual participant data cannot be provided. |
| 8.4 | Source code access | No | https://github.com/michael-nakai/waterway |
| 8.5 | Full results | Yes | Page 12-15 |
| **Results** | | | |
| 9 | Descriptive data | Yes | Page 1-15 |
| 10 | Microbiome data | Yes | Page 14-15 |
| 10.1 | Taxonomy | Yes | Page 15 |
| 10.2 | Differential abundance | Yes | Page 14 |
| 10.3 | Other data types | Yes | Page 15 |
| 10.4 | Other statistical analysis | Yes | Page 15 |
| **Discussion** | | | |
| 11 | Key results | Yes | Page 15-16 |
| 12 | Interpretation | Yes | Page 16-17 |
| 13 | Limitations | Yes | Page 17-18 |
| 13.1 | Bias | NA | |
| 13.2 | Generalizability | NA | |
| 14 | Ongoing/future work | Yes | Page 17-18 |
| **Other information** | | | |
| 15 | Funding | Yes | Page 18 |
| 15.1 | Acknowledgements | Yes | Page 18 |
| 15.2 | Conflicts of Interest | Yes | Page 19 |
| 16 | Supplements | NA | |
| 17 | Supplementary data | NA | |

**Table S4.** Comparison between baseline randomisation arms.

| Trait | Diet A (n=13) Mean ± SD | Diet B (n=7) Mean ± SD | *P*-value |
|---|---|---|---|
| Age | 57 ±11.23 | 53.57 ±12.58 | 0.14 |
| Body mass index | 25.05 ±2.64 | 26.99 ±1.98 | 0.67 |
| Creatinine | 79.62 ±14.69 | 78.14 ±8.09 | 0.81 |
| eGFR | 81.54 ±10.37 | 89.71±0.49 | 0.05 |
| Glucose | 5.15 ±1.09 | 5.07 ±0.74 | 0.87 |
| Total protein | 75.31 ±4.23 | 75.71 ±5.06 | 0.85 |
| Albumin | 45.23 ±9.17 | 42.71 ±3.86 | 0.50 |
| Globulin | 32.92 ±3.52 | 33.0 ±3.56 | 0.96 |
| Bilirubin | 13.69 ±6.31 | 14.85 ±4.41 | 0.67 |
| ALT | 22.08 ±9.33 | 28.28 ±10.77 | 0.19 |
| GGT | 27.31 ±27.17 | 27.29 ±10.36 | 0.99 |
| ALP | 59.92 ±24.83 | 61.00 ±9.27 | 0.91 |
| Cholesterol | 4.95 ±0.97 | 6.06 ±1.38 | 0.05 |
| Cholesterol HDL | 1.54 ±0.43 | 1.33±0.38 | 0.30 |
| Cholesterol non-HDL | 3.42 ±0.98 | 4.73±1.28 | **0.02** |
| Cholesterol LDL | 2.92 ±0.928 | 3.99 ±1.18 | **0.04** |
| Triglycerides | 1.08 ±0.34 | 1.59 ±0.62 | **0.03** |
| Office SBP | 139.83 ±12.85 | 148.14 ±11.25 | 0.17 |
| Office DBP | 84.13 ±10.75 | 80.93 ±8.71 | 0.51 |
| Office HR | 68.23 ±11.41 | 63.71 ±16.26 | 0.48 |
| 24h SBP | 135.38 ±6.53 | 137 ±3.51 | 0.55 |
| 24h DBP | 87.00 ±7.34 | 88.00 ±6.03 | 0.76 |
| 24h HR | 69.62 ±8.48 | 64.29 ±7.87 | 0.19 |

Legend: significance highlighted in bold. eGFR, estimated Glomerular Filtration Rate; ALT, alanine aminotransferase; GGT, gamma-glutamyl transferase; ALP, alkaline phosphatase; HDL, high-density lipoprotein; LDL, low-density lipoprotein; SBP, systolic blood pressure; DBP, diastolic blood pressure; 24h, 24 hours. n=20/group.

**Table S5.** Comparison of changes in biochemical parameters from baseline in the placebo and HAMSAB diet arm.

| Characteristic | Baseline | Placebo | *P*-value | Baseline | HAMSAB | *P*-value |
|---|---|---|---|---|---|---|
| Body mass index | 25.7±2.5 | 25.7±2.6 | 0.42 | 25.8±2.6 | 25.8±2.5 | 0.53 |
| Creatinine | 80.1±13.1 | 80.5±15.0 | 0.43 | 79.1±12.9 | 77.9±10.1 | 0.79 |
| eGFR | 84.3±9.8 | 85.5±6.5 | 0.34 | 83.3±9.7 | 83±11.1 | 0.74 |
| Glucose | 5.0±0.6 | 5.3±1.2 | 0.33 | 5.2±1.0 | 5.2±0.8 | 0.74 |
| Total protein | 75.3±4.6 | 75.7±3.3 | 0.43 | 75.5±4.5 | 74.7±3.8 | 0.38 |
| Albumin | 42.3±3.5 | 41.7±3.0 | 0.62 | 44.2±7.7 | 41.7±2.7 | 0.22 |
| Globulin | 33.0±3.9 | 34.0±3.0 | 0.35 | 33.2±3.2 | 33±3.1 | 0.80 |
| Bilirubin | 13.7±6.0 | 13.7±4.8 | 1.00 | 13.9±5.5 | 12.7±4.5 | **0.04** |
| ALT | 25.0±10.4 | 24.2±10.5 | 1.00 | 24.6±9.8 | 28.3±15.7 | 0.10 |
| GGT | 29.6±32.2 | 30.2±28.2 | 0.63 | 27.8±22.2 | 30.9±26.4 | 0.05 |
| ALP | 61.6±19.0 | 62.4±20.3 | 0.63 | 60.7±20.7 | 61.9±19.8 | 0.35 |
| Cholesterol | 5.5±1.5 | 5.6±1.3 | 0.51 | 5.3±1.0 | 5.6±1.2 | 0.06 |
| Cholesterol HDL | 1.5±0.4 | 1.5±0.3 | 0.73 | 1.5±0.4 | 1.5±0.4 | 1.00 |
| Cholesterol non-HDL | 4.1±1.5 | 4.1±1.3 | 0.72 | 3.9±1.1 | 4.2±1.2 | **0.02** |
| Cholesterol LDL | 3.5±1.3 | 3.4±1.2 | 0.66 | 3.3±1.0 | 3.5±1.1 | **0.03** |
| Triglycerides | 1.3±0.6 | 1.3±0.7 | 0.80 | 1.3±0.8 | 1.5±0.9 | **0.01** |

Legend: significance highlighted in bold. eGFR, estimated Glomerular Filtration Rate; ALT, alanine aminotransferase; GGT, gamma-glutamyl transferase; ALP, alkaline phosphatase; HDL, high-density lipoprotein; LDL, low-density lipoprotein. n=20/group. *P*-value shows comparison between groups (baseline vs placebo, or baseline vs HAMSAB).

**Table S6.** Summary of gastrointestinal symptoms based on the 100mm Visual Analogue Scale responses for stool habits.

| Symptoms | Baseline | Placebo | *P*-value | Number of pairs | Baseline | HAMSAB | *P*-value | Number of pairs |
|---|---|---|---|---|---|---|---|---|
| Abdominal symptoms (mm) | 18.86±22.45 | 13.36±17.86 | 0.370 | 14 | 15.54±18.02 | 26.31±24.64 | 0.123 | 13 |
| Abdominal pain (mm) | 14.21±15.11 | 10.93±12.27 | 0.253 | 14 | 11.08±11.86 | 21.15±18.00 | **0.024** | 13 |
| Bloating (mm) | 25.64±26.22 | 16.79±19.26 | **0.022** | 14 | 18.31±24.72 | 26.62±28.68 | 0.270 | 13 |
| Wind (mm) | 35.29±30.01 | 23.00±25.94 | 0.064 | 14 | 27.25±30.54 | 26.42±28.50 | 0.895 | 13 |
| Tiredness/lethargy (mm) | 25.07±19.41 | 15.64±13.80 | 0.110 | 14 | 23.54±20.59 | 32.23±26.14 | 0.240 | 13 |
| Nausea (mm) | 16.36±24.38 | 8.00±9.46 | 0.234 | 14 | 10.77±12.71 | 13.77±15.78 | 0.350 | 13 |

Legend: data shown as mean±SD. Symptoms were measured out of 100 mm. No significant differences in baseline responses were found between the two arms.

**Table S7.** Summary of the results for 24-h day and night blood pressure.

| Parameter | Baseline | Placebo | *P*-value | Baseline | HAMSAB | *P*-value |
|---|---|---|---|---|---|---|
| Day SBP | 139.7±11.6 | 140.2±10.8 | 0.82 | 142.1±10.6 | 138.1±10.3 | **0.043** |
| Day DBP | 90.5±7.0 | 90.5±8.1 | 1.00 | 91.6±7.9 | 90.2±8.1 | 0.25 |
| Night SBP | 120.45±9 | 121.5±12.2 | 0.66 | 123.9±10.2 | 119.1±11.4 | **0.02** |
| Night DBP | 75.3±8.29 | 75.57±11.12 | 0.90 | 76.3±9.2 | 74.1±9.24 | 0.08 |

Legend: significance highlighted in bold. SBP, systolic blood pressure; DBP, diastolic blood pressure.

**Table S8.** Taxonomic changes in the faecal microbiome between placebo and HAMSAB.

| **Taxonomic changes** | **log2FC** | **logCPM** | ***P*-value** | **FDR q-value** |
|---|---|---|---|---|
| Pasteurellaceae | -5.78 | 12.196 | 3.33E-06 | 0.00059 |
| *Azospirillum*_sp__47_25 | -4.28 | 11.270 | 2.73E-05 | 0.00167 |
| *Ruminiclostridium*_5 | -3.38 | 10.705 | 2.85E-05 | 0.00167 |
| *Ruminococcus_gauvreauii*_group | -3.52 | 12.951 | 6.50E-05 | 0.00286 |
| *Parabacteroides_distasonis* | -2.82 | 16.701 | 9.44E-05 | 0.00332 |
| bacterium_YE57 | -3.21 | 11.037 | 0.000549 | 0.01611 |

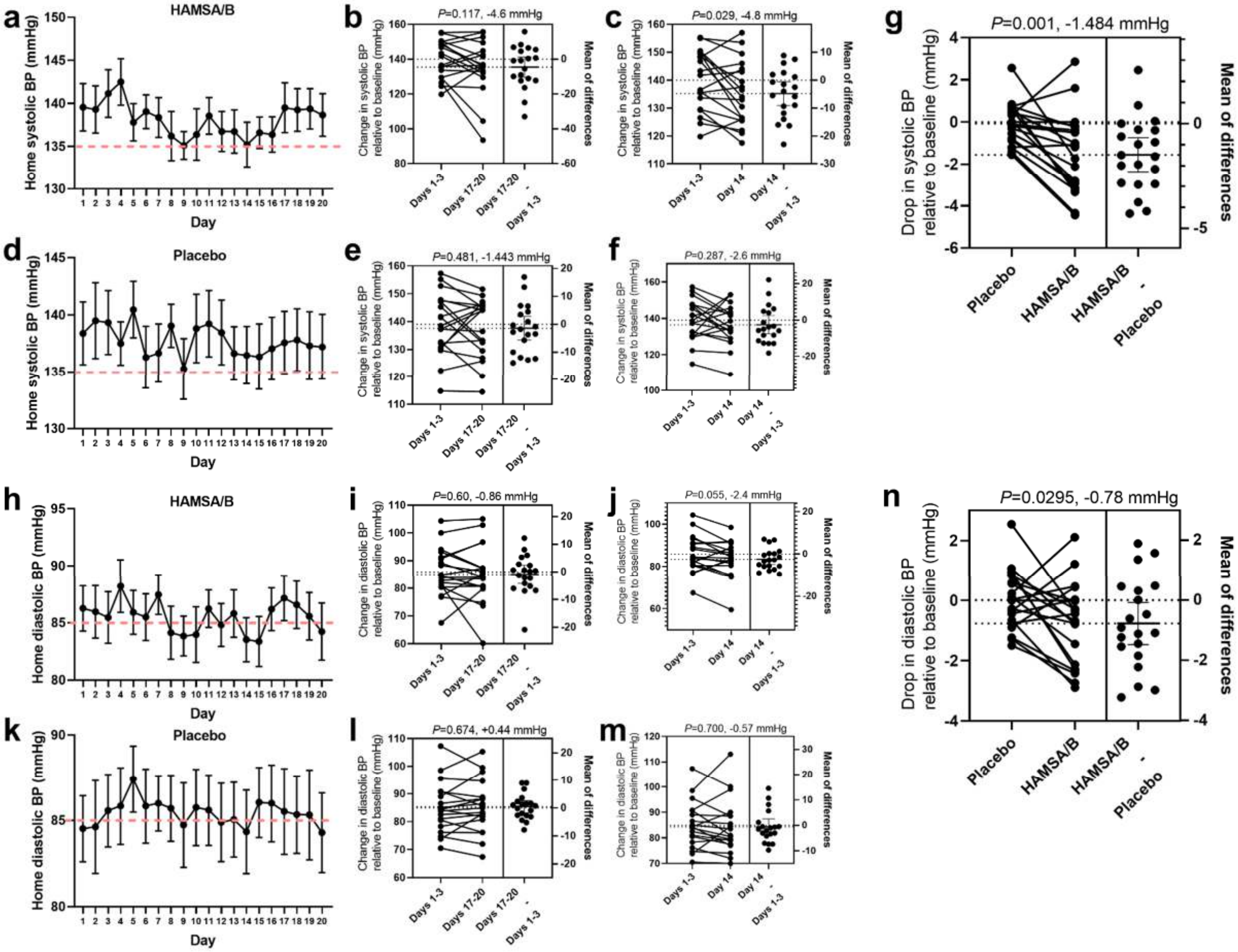
a
HAMSA/B
Home systolic BP (mmHg)
Day
b
P=0.117, -4.6 mmHg
c
P=0.029, -4.8 mmHg
g
P=0.001, -1.484 mmHg
Drop in systolic BP relative to baseline (mmHg)
Mean of differences
Placebo
HAMSA/B
HAMSA/B - Placebo
d
Placebo
e
P=0.481, -1.443 mmHg
f
P=0.287, -2.6 mmHg
Change in systolic BP relative to baseline (mmHg)
Days 1-3
Days 17-20
Days 17-20 - Days 1-3
Day 14
Day 14 - Days 1-3
h
HAMSA/B
Home diastolic BP (mmHg)
i
P=0.60, -0.86 mmHg
j
P=0.055, -2.4 mmHg
n
P=0.0295, -0.78 mmHg
Drop in diastolic BP relative to baseline (mmHg)
k
Placebo
l
P=0.674, +0.44 mmHg
m
P=0.700, -0.57 mmHg
Change in diastolic BP relative to baseline (mmHg)

**Figure S1: HAMSB diet reduces in home systolic and diastolic blood pressure (BP)**. a) mean home systolic BP over 21-day HAMSAB intervention. b) mean change in systolic BP relative to baseline and the mean difference in HAMSAB treated participants at b) Day1-3 and Day 17-20 and c) Day 1-3 and Day 14. d) mean change in systolic BP relative to baseline and the mean difference in placebo treated participants at e) Day1-3 and Day 17-20 and f) Day 1-3 and Day 14. g) overall drop in systolic BP relative to baseline and placebo-subtracted mean difference. h) mean home diastolic BP over 21-day HAMSAB intervention. mean change in diastolic BP relative to baseline and the mean difference in HAMSAB treated participants at i) Day1-3 and Day 17-20 and j) Day 1-3 and Day 14. k) mean change in diastolic BP relative to baseline and the mean difference in placebo treated participants at l) Day1-3 and Day 17-20 and m) Day 1-3 and Day 14. n) overall drop in diastolic BP relative to baseline and placebo-subtracted mean difference. n=20/treatment group. Error bars represent ±SEM.

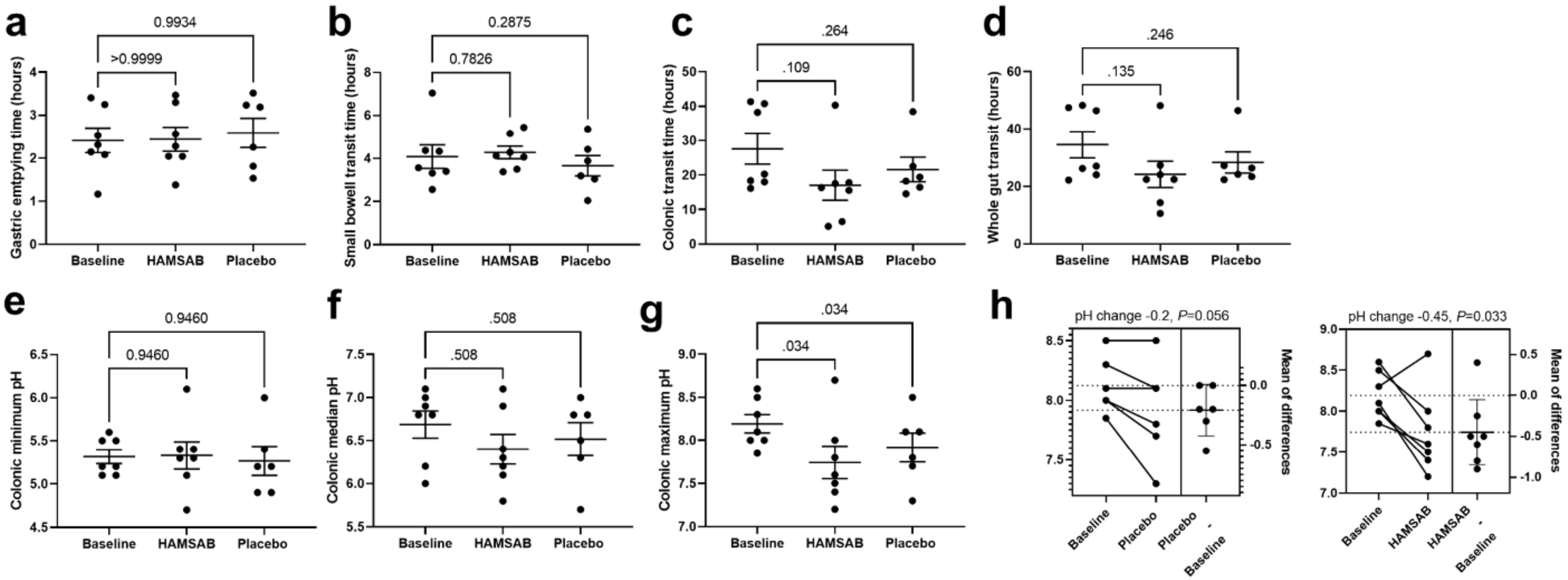


**Figure S2: The impact of HAMSAB diet on gastrointestinal pH and transit times measured by Smart Pill.** We observed no difference in a) gastric emptying time, b) small intestinal transit time, c) colonic transit, and d) whole gut transit time in hours. We observed no difference in colonic e) minimum and f) median pH, but observed a decrease in g) maximum pH. h) Summary of maximum colonic pH in placebo and HAMSAB relative to baseline. Placebo data from a participant who had antibiotics between visits 3 and 4 was removed, resulting in n=6 for placebo and n=7 for baseline and HAMSAB groups. One-way ANOVA adjusted for multiple comparisons, showing adjusted P-values. Error bars represent mean±SEM.

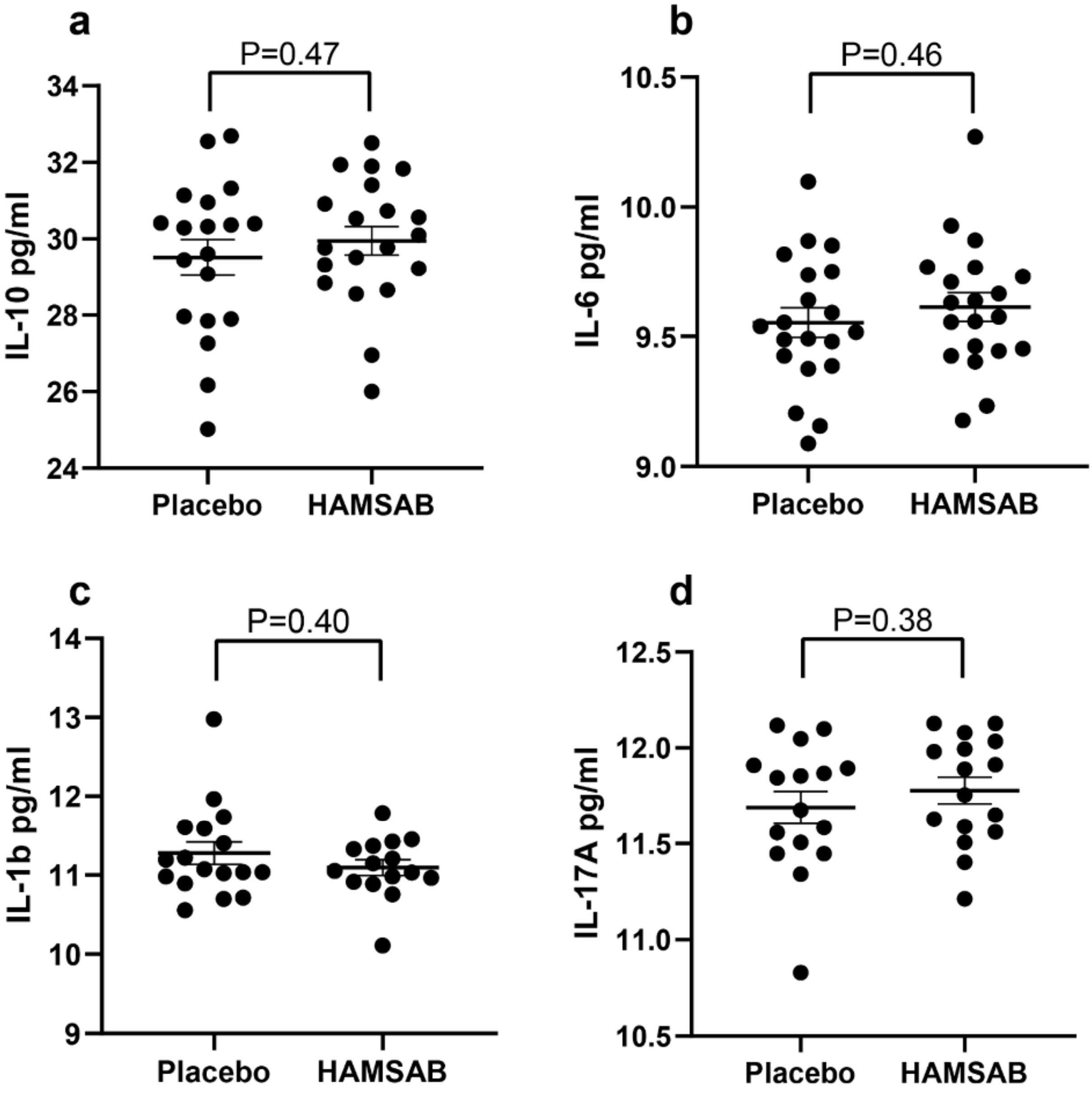


**Figure S3: HAMSAB diet does not alter plasma cytokine levels over placebo.** Plasma levels of a) IL10, b) IL-6, c) IL-1β and d) IL-17A in placebo and HAMSAB treated participants. n=16-20/treatment group. Error bars represent mean±SEM.
